# Data quality and associations over 4 years of use of WHO’s Clinical Registry - Trauma Module, in Rwanda

**DOI:** 10.64898/2026.01.24.26344755

**Authors:** Indrakantha Welgama, Jean Nepomuscene Ntezimana, Irene Bagahirwa, Venantie Umuhoza, Fabrice Iradukunda, Daniel Lange, Leila Ghalichi, Sudha Jayaraman, Kathryn Chu, Aurore Nishimwe, Ephrem Daniel Sheferaw, Faustin Ntirenganya, Jean Claude Byiringiro, Justine Davies

## Abstract

**Objective:** Poor quality of injury care causes avoidable death and disability. Quality data is integral to care-quality improvement. We assessed longitudinal data quality of the Trauma Module (TM) of WHO’s Clinical Registry in Rwanda, towards understanding data collection challenges in low resourced settings.

**Methods:** Retrospective analysis of TM data between its implementation in August 2019 and July 2023. We describe trends in overall TM case and variable missingness and their associations with availability of funding and formal training. Variable missingness is also described by each of the 7 domains of the TM.

**Findings:** The TM was initiated in 4 hospitals in August 2019 and expanded to 6 in June 2021. Between implementation and July 2023, 13,513 cases were entered. Annual numbers of cases varied per hospital (maximum of 2051, minimum of 259). Case missingness ranged from 60% to 66% across all hospitals. The median annual variable missingness varied between hospitals from 28% to 41%. Over the whole period, across all hospitals, median missingness was 39.63% (IQR 0.65 – 82.46); 12 variables had 100% missingness and 15 variables, 0% missingness. Variable missingness depending on TM domain; highest in “In-ward care & Facility disposition” (77.30%) and “EU disposition” (51.67%). Data collection was dependent on external funding; when funding ceased in 2023, case and variable missingness increased to 100%. Short term increases in data collection were noted following periodic formal trainings.

**Conclusions:** Although TM sustainment in Rwanda relied on external funding, even with funding and training, data quality was variable.

## Introduction

Intentional and unintentional injuries result in approximately 8% of global deaths [1]. Over 90% of the global injury burden is experienced by Low-or-Middle Income Countries (LMICs) [2]. It is estimated that 60% of the burden of avoidable post-injury deaths could be averted with better care at health facilities [3].

The United Nations (UN) Sustainable Development Goal 3 (UN SDG 3) identifies “Good Health and Wellbeing” as an important component of global development [4]; the UN has mandated that all member states provide citizens with good quality healthcare service [5]. A 2018 report by the World Health Organization (WHO) and World Bank (WB) highlighted poor quality health services as a key factor slowing down progress on improving healthcare in all countries [6]. Quality data - which are complete and accurate – are essential for development and sustainment of high-quality healthcare services [2, 7-9] and reducing injury-related morbidity and mortality.

Many LMICs have systems for collecting patient data, such as the Indoor Morbidity and Mortality Records (IMMR) and hospital patient admission registers [10]. Some of these have progressed from paper-based to digitalized databases, such as the District Health Information Software 2 (DHIS2) system [11]. Although a comprehensive electronic health record system is not feasible in many LMICs, many utilise condition-specific databases or registries based on information needs [12, 13]. Injury registries, which collect reliable and usable data, are a potential means to inform injury care quality improvement [14, 15].

The World Health Assembly (WHA), through resolution 72.16 (2019) called on countries “*to implement mechanisms for standardized data collection to characterize the local acute disease burden and identify high-yield mechanisms for improving the coordination, safety and quality of emergency care worldwide*” [16]. In response to this and to support the emergency and injury care quality improvement needs of its member states, the WHO developed the International Registry for Trauma and Emergency Care (IRTEC) in 2019 [17]. IRTEC has since expanded to the WHO Clinical Registry by including modules on primary, emergency, critical and operative care. The injury care module of the WHO Clinical Registry, hereafter referred to as the “Trauma Module (TM)” remains the most widely implemented application; it is an online database, using the DHIS2 data entry platform [18]. It is maintained by WHO, with implementing states having ownership of their data. The TM captures patient related, prehospital, prior facility, Emergency Department (ED), and in-ward data, as well as outcomes of injured patients [17, 18]. It consists of a mandatory core (251 questions) and an extended dataset (23 questions), with a total of 274 questions, to produce 151 output variables. These are categorized under 7 domains (Table 1). Standard visualisations allow quick interpretation of data for use in clinical quality improvement. The TM is currently used by several institutions in different countries [19].

**Table 1.** Sections of the WHO Clinical Registry (Trauma Module) and their numbers of output variables.

|  | **Domain** | **No. of Variables** |
| --- | --- | --- |
| 1 | Registry & Demography | 20 |
| 2 | Pre-Emergency Department care | 22 |
| 3 | Context of the injury | 31 |
| 4 | Details of the injury | 19 |
| 5 | Care in the Emergency Department | 27 |
| 6 | Emergency Department Disposition | 10 |
| 7 | In-ward Care & Facility Disposition | 22 |

The Republic of Rwanda, a LMIC in East Africa, implemented the complete TM in 2019 as a step towards country-wide injury surveillance, initiated by the Rwanda Biomedical Centre (RBC), under the guidance of the Ministry of Health. Data were collected at secondary and tertiary level hospitals by data collectors (usually ED nurses) onto paper-based forms; later entered into the electronic database by a data entry officer. Rwanda is one of the few LMICs which has attempted to ensure continuous data collection into the TM and although some challenges in implementation have been previously described in other settings [14, 20, 21], very few studies have assessed quality of data over time, and none that we are aware has assessed data quality over multiple years [22]. To anticipate and address future registry implementation challenges, we assessed long-term data quality in Rwanda’s TM and associate this with training and funding.

## Methods

### Study Design

A retrospective analysis of secondary data from the TM of the WHO Clinical Registry from 01^st^ August 2019 to 31^st^ July 2023.

### Study setting and description of data

The TM implementation was initially funded by the United States National Institute of Health (US NIH); funds were for implementation of the registry at 4 hospitals, supporting basic equipment, data collection on all injury patients presenting to the emergency departments (ED) (via a financial incentive to data collectors of 216,000 RWF per month), and training, with follow up for 2 years. Two hospitals were added in June 2021 and by the end of the funding in August 2021, 6 hospitals were collecting data: two within the urban settings of the capital city, Kigali, and four within each one of the 4 provincial regions. Following the end of US NIH funding, the WHO assisted RBC with funding from the African Federation for Emergency Medicine (AFEM) for continuation of the TM for a further year, until September 2022, after-which all external funding ended, although data collectors were still expected to input data.

### Data Collection and Analysis

Anonymised data were downloaded from 01^st^ August 2019 to 31^st^ July 2023. These were cleaned and analysed using STATA 18.0 (StataCorp LLC, USA). Data are described using numbers and percentages, and measure of central tendency and spread, depending on distribution.

Data quality is presented as percentage case missingness or variable missingness. We had no access to case notes, hence could not assess data accuracy. Case missingness is defined as the number of patients with injury who were not entered into the TM during a given period, as a proportion of the total number of injured patients reported as seen in the ED. The latter was calculated from the Hospital Management Information System (HMIS). i.e. *case missingness = (number of ED admission cases for injury captured into HMIS – number of cases entered into the TM)/number of ED admission cases for injury capture into HMIS*. The TM and the HMIS are two separate digital data entry systems.

Variable missingness is presented as the average of percentage missingness of each variable for each case entered into the TM, accounting for skip patterns. The annual percentage variable missingness is presented for all hospitals and variables; variable missingness is also presented for each TM domain.

### Ethics Approval

This analysis was done as part of two larger studies which received ethics approval from Rwanda National Research Ethics Committee of the Ministry of Health, Rwanda (ERC No. 85/RNEC/2023, No 14/RNEC/2024, and No.99/RNEC/2023) and the ethics committee of Kigali University Teaching Hospital, Rwanda (EC/CHUK/036/2023).

## Results

The TM in Rwanda was implemented in 3 tertiary level hospitals and 3 secondary level hospitals (Table 2).

**Table 2.** Description of the Hospitals where Trauma Module is implemented.

| Hospital | Facility Level | Setting | Commencement of Trauma Module Data collection |
| --- | --- | --- | --- |
| Hospital 1 | Tertiary Level Care | Urban – Kigali | August 2019 |
| Hospital 2 | Tertiary Level Care | Urban - Provincial | August 2019 |
| Hospital 3 | Tertiary Level Care | Urban – Kigali | August 2019 |
| Hospital 4 | Secondary Level Referral Hospital | Urban - Provincial | August 2019 |
| Hospital 5 | Secondary Level Referral Hospital | Urban - Provincial | June 2021 |
| Hospital 6 | Secondary Level Referral Hospital | Urban - Provincial | July 2022 |

A total of 13,513 cases were entered into the TM. The median number of cases entered monthly was 326 (IQR 179-400) with the highest number entered in November 2020 (553 cases), while no cases were entered in April, May and June 2023 (Table 3). In 2023, only 2 hospitals entered any data.

**Table 3.**
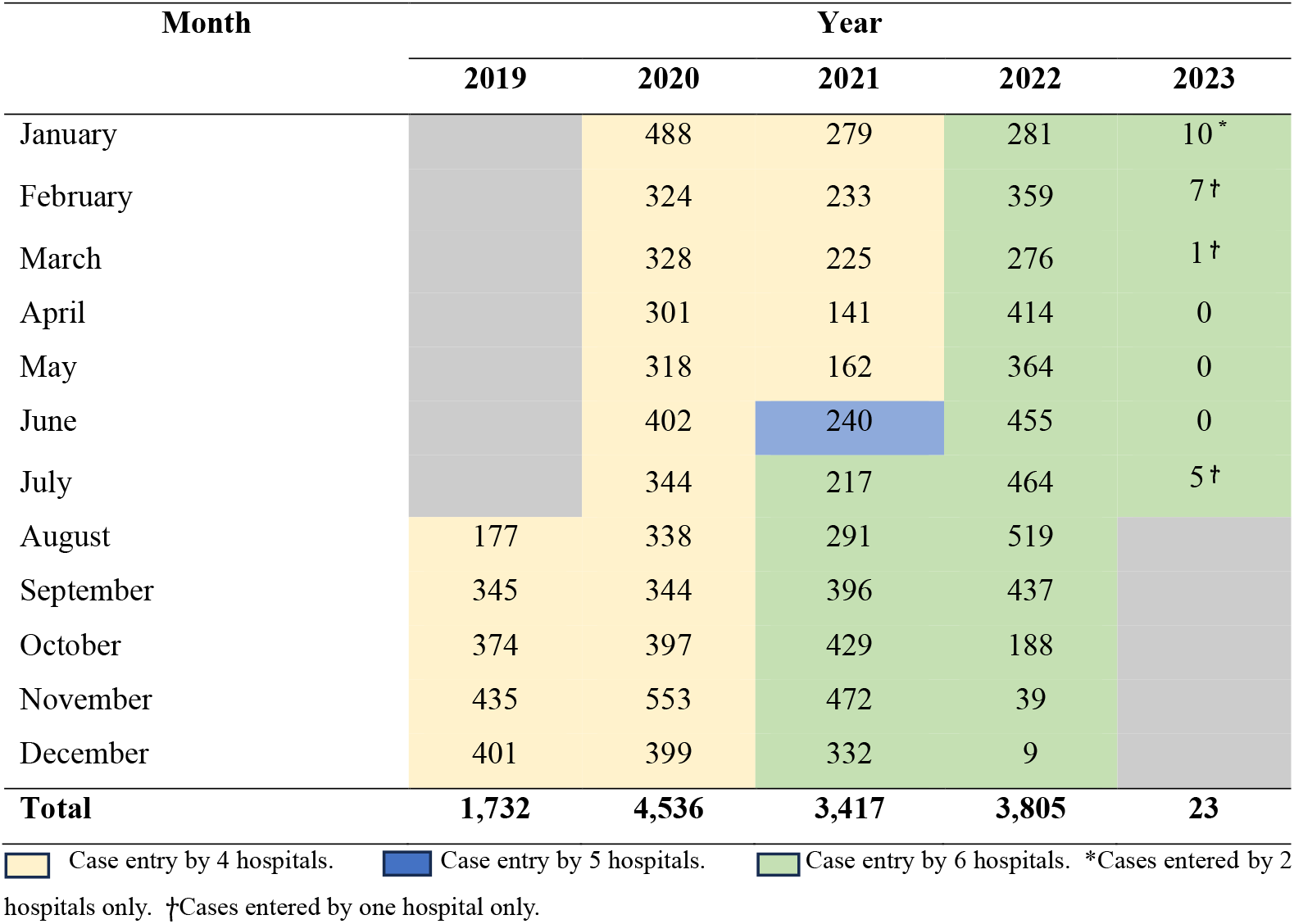
Trauma Module Monthly Case Entries.

Annual case entry by each hospital into the TM are shown in Table 4. Hospital 1 entered highest number of cases, whereas Hospitals 5 and 6, both of which started data collection later, had entered fewer cases.

**Table 4.** Annual Case Entry into the Trauma Module by Hospital.

| Hospital | Year |  |  |  |  | Total<br>(n=13,513) |
| --- | --- | --- | --- | --- | --- | --- |
|  | 2019 | 2020 | 2021 | 2022 | 2023 |  |
| Hospital 1 | 803 | 2,051 | 1,031 | 832 | 0 | 4,717 |
| Hospital 2 | 352 | 961 | 822 | 549 | 15 | 2,699 |
| Hospital 3 | 318 | 819 | 527 | 547 | 0 | 2,211 |
| Hospital 4 | 259 | 705 | 527 | 611 | 8 | 2,110 |
| Hospital 5 | NR* | NR* | 334 | 566 | 0 | 900 |
| Hospital 6 | NR* | NR* | 176 | 700 | 0 | 876 |

\*NR – Not Reported as TM not implemented.

Numbers of cases entered per month steadily increased during the first 4 months and thereafter fluctuated, with a prominent reduction seen during mid-2021 (Figure 1).

**Figure 1.**
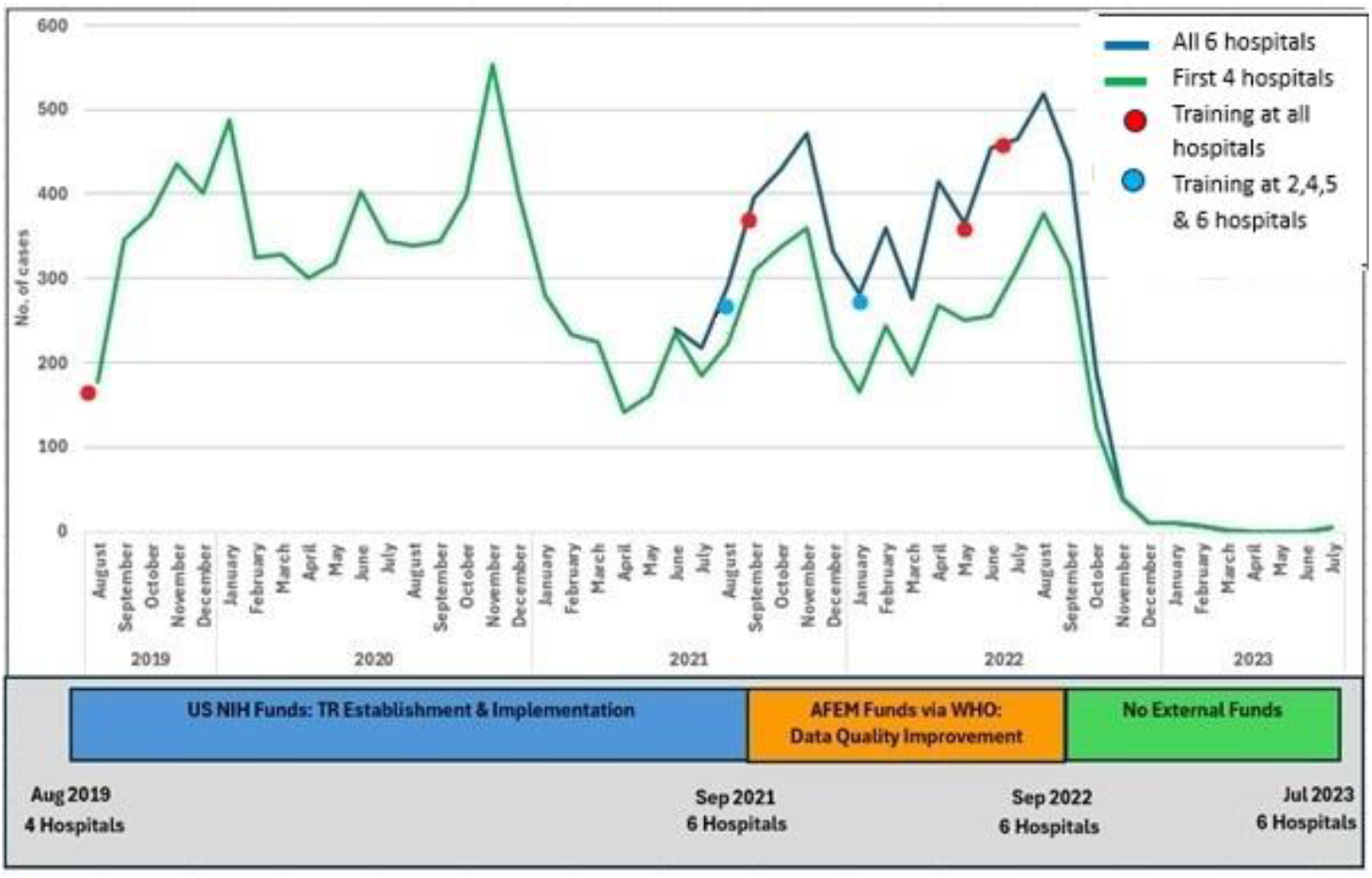
Trauma Module Case Entry (Aug 2019 – Jul 2023). Points indicate when training was given. Funding sources and duration are shown at the bottom of the figure.

Case entry into the TM appears to vary with the availability of external funding, with peaks observed during the initial phases of funding by US NIH and by AFEM. When external funding ceased after September 2022, data entry rapidly declined and by April 2023, it had halted in all 6 hospitals (Figure 1).

Six formal training sessions were conducted between August 2019 to July 2023 (Supplementary material – Appendix 1). Initial training was conducted in June 2019 prior to the implementation of the TM, and thereafter no further formal training was conducted over the next 2 years, although informal mentoring sessions were conducted at individual hospitals, as deemed necessary. During the AFEM funding period, 5 formal training sessions were conducted. Following each training, an uptick in the numbers of cases entered into the TM is observed (Figure 1). Although there were no formal processes in place for audit and feedback of the quality of data entry into the TM, there were ad-hoc communications and occasional visits to the sites by RBC team, to inquire and advice on the numerous challenges encountered in data collection. An attempt at assessing the data quality in February 2020 was foiled by COVID-19 restrictions.

Injury specific HMIS data were unavailable for 2019 and 2020. For 2021 and 2022 a total of 8,155 and 5,822 injury cases were recorded into HMIS at all 6 hospitals, respectively. Giving an annual case missingness of 58.10% and 36.76% for 2021 and 2022, respectively (Table). Given technical failures of data entry into HMIS during the study period and the lack of information on the duration of the technical failures, we did a sensitivity analysis excluding hospitals with HMIS failures for the entirety of the year in which the failures occurred; this gave annual case missingness rates of 66.32% and 60.06% in 2021 and 2022, respectively (Table 5). Case missingness for 2023 was not calculated as TM monthly case reporting was mostly fewer than 10 cases (Table 3).

**Table 5.**
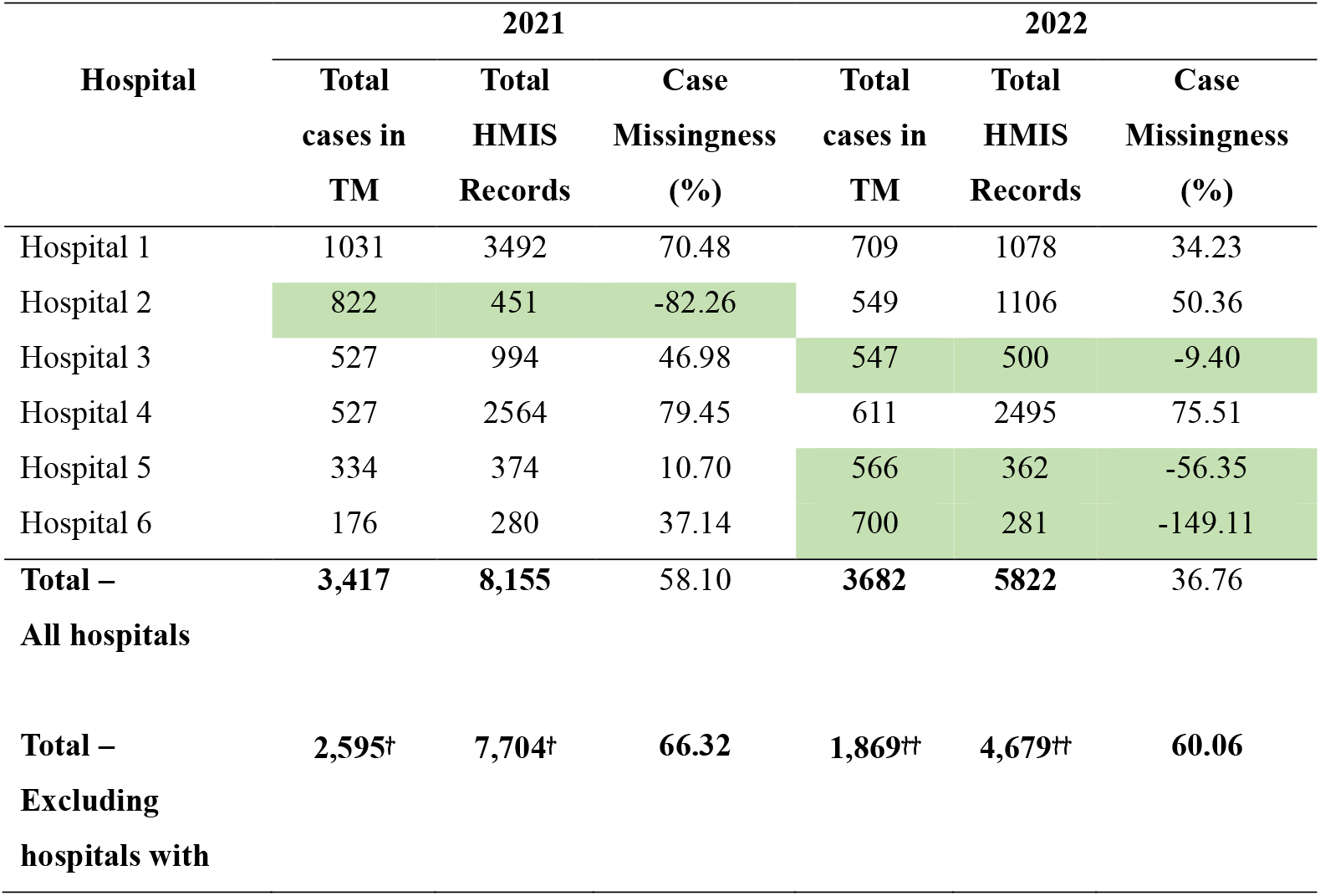
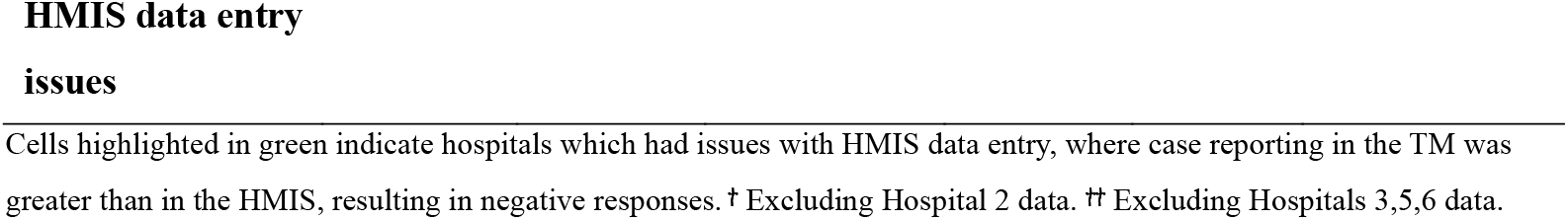
Annual Percentage Case Missingness 2021-2022.

Over the whole time period, across all hospitals, there were 12 variables with 100% missingness and 15 variables with 0% missingness; some variables such as the “Organisation unit”, “Date of arrival to the ED” were always complete (0% variable missingness), while some variables such as “Patient arrival hour”, “Use of substance - drugs” were always empty (100% variable missingness), in all hospitals throughout the period. Median missingness was 39.63% (IQR 0.65 – 82.46). The median annual variable missingness varied between hospitals from 28% and 41% (Figure 2). Among individual hospitals, hospital 6 had the highest annual variable missingness, while hospitals 1 and 4 had an increasing trajectory of missingness over time.

**Figure 2.**
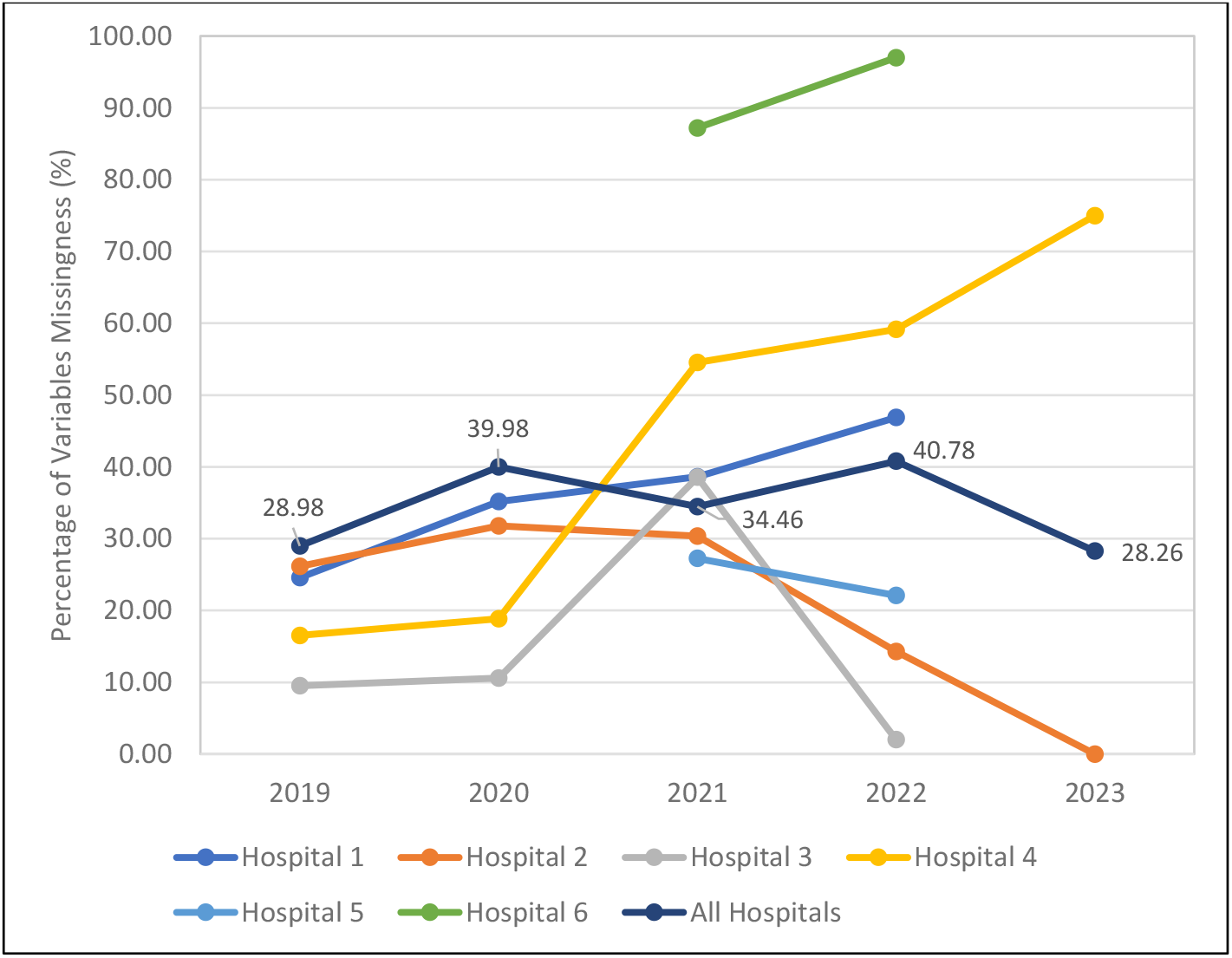
Annual Median Percentage Variables Missingness 2019 to 2023 (Overall and individual hospitals). Data values represent the points of the “All Hospitals” Percentage Variables Missingness line

The highest median percentage variable missingness was observed in the domain of In-ward Care & Facility Disposition (77.3%), while the lowest median percentage variables missingness was observed in the domain of Injury Details (26.5%) (Supplementary material – Appendix 2).

## Discussion

Missingness of cases and variables for the 251 questions in the core module of the WHO TM implemented in Rwanda were often substantial, although this improved following periodic training. Variables where information was available from the ED such as the type and mechanism of injuries were captured reasonably well. However, data on patient outcomes, a key variable for understanding effectiveness of care delivery, had the highest variable missingness. This is likely because these outcomes are not readily available to ED-based data collectors, and they require follow up of patients or access to inpatient records. Data inputted were rarely, if ever, used for healthcare quality improvement purposes, and healthcare staff didn’t articulate their utility for healthcare quality improvement. Limited ownership of the TM was perceived at the hospital level. Importantly, TM sustainability in Rwanda was entirely dependent on external funding, despite the presence of a dedicated in-country team at RBC consisting of a part time manager and data oversight assistant.

Poor data quality challenges its utility for service delivery or planning, potentially compromising patient outcomes [22, 23]. In addition to facilitating care quality improvement, understanding barriers to improving data quality are important to reduce waste; given the implementation and maintenance of a registry incurs substantial cost, time, and effort [24]. This is especially important in low resourced settings where opportunity costs of investment are substantial. However, that issues in data quality were sustained over a long period of time in a country which aspires to utilize data to inform health care [25] and has a high burden of injuries [26] and injury deaths - which are targeted for action by Ministry of Health - provides opportunities for learning for other low resourced countries looking to implement such registries. Indeed, given the universality of data-quality issues, lessons learnt from Rwanda likely have broad applicability regardless of country income level.

On the surface, according to the COM-B model [27], the data collectors were capable (they were trained), they had opportunity (the TM was ready-designed and equipment was supplied), and they had some motivation (they were paid an incentive during periods of implementation). However, behaviour towards ensuring high quality of data was not optimised. Whilst these proximate mediators of quality data collection might be improved upon, we argue that there are several larger conditions which need to be considered before such registries are likely to capture data of high enough quality for meaningful healthcare quality improvement.

First, the impetus to deliver data-informed healthcare quality improvement needs to be clearly articulated by local healthcare providers, policy makers, or planners prior to data collection. Whilst data are necessary for quality improvement initiatives, the presence of data alone is not sufficient to drive these. It is essential that the data collectors as well as the hospital-level facilitators and administrators have a desire to utilise data, are aware of the importance of inputting high quality data into registries and understand the adverse outcomes that may occur in the case of poor-quality data collection. The lack of data usage by the hospitals here may indicate the need for more formal trauma quality improvement training which has not been available in Rwanda. However, a pilot was conducted recently, and results are being evaluated for publication.

Second, variables should be selected based upon necessity for delivering quality improvement and the feasibility of their collection. Where possible, data collection into a registry should also be integrated within the existing data infrastructure in a health system. The core module of the WHO TM contained 251 variables, which are usually to be collected by busy clinicians in healthcare facilities which often struggle to provide clinical care.[28] The competing priorities experienced by an under-resourced health workforce, both drive and conspire with a lack of prioritisation of record keeping in many low resourced settings to challenge good clinical note taking.[14, 19] Indeed, in our work in Rwanda, we found that even data recording into the HMIS system used for official government-level reporting for service delivery and planning was unreliable and without good data governance structures. On informal exploration, we also found that although there is a move towards electronic health records in Rwanda, these suffer similar issues of variable completeness to the TM, especially in those variables concerning patient outcomes or disposition. The lack of availability of data for clinical care and outcomes in paper-based notes or electronic records means that to complete data entry for the TM data needs to be specifically collected for this purpose. This poses problems with data quality when patients move to in-patient wards where the care-focus is not specifically trauma, as we have shown. The lack of inter-operability among different data collection systems existing within a facility compounds these issues, leading to multiplication of effort and further waste.

Third, implementation of data governance structures, including Standard Operational Procedures (SOPs) for data collection in combination with regular data quality checks and feedback to staff, is needed to ensure that data collected is of high quality. Prior to our assessment, there had been no formal data quality checks, such that neither the staff collecting nor appraising the data were aware of any issues. Whereas data quality checks as part of audit and feedback cycles have been shown to improve quality both within and outside of research.[29]

Fourth, the feasibility of sustainable financing for data collection should be considered prior to commencing data collection into a registry. Implementation and sustainment of registries have been described as financially challenging in most LMIC settings, and dependent on the support of external funding sources [14, 20]. This was evident in our study, given that data collection effectively ceased with cessation of external funding, with no financial support available from the government to sustain the TM. The cost of maintaining the TM in Rwanda is mainly related to the routine data capturing process including the resources and connectivity, training, and administrative expenses, similar to descriptions in other studies [14, 15, 21]. Monthly stipends for data collectors have been recommended as a financial incentive in prior publications [14, 21], however, we found these financial incentives are not adequate to ensure high quality data collection. Although controversial, pay for performance may possibly increase data quality, though this is unlikely to be sustainable in the LMIC setting [30].

Fifth, making data collection mandatory – in combination with robust quality assurance systems [7, 8, 21-23] – is likely to substantially improve data collection. For example, the UK’s National Major Trauma Registry [31] has over 600 variables, the collection of which is mandated by government. Accompanying this mandate is funding, good data governance structures, and staff dedicated to running the registry. Whereas such investment is unlikely to be achievable in low-income countries, especially in the current funding environment. Nevertheless, lessons in maintaining registries in low resource settings can be learnt from a successful example of a registry for critical care patients across Africa and Asia, the Critical Care Africa Asia (CCAA) registry. Although developed for LMICs using funding from the UK’s “Wellcome Trust”, data-quality in the registry is high [32] and sustained over the long term, with a decentralised leadership structure. The CCAA meets several of the conditions which we have outlined as required for success. It was co-developed by future data users based on their needs for data use for clinical care quality improvement and research. Both training and implementation methods were used to embed data collection and use into routine practice, and the potential for local users to use the networked registry as a foundation for trials and other research forms an extra incentive.[33] Another important consideration is that the CCAA was established on already fertile ground; the requirement for understanding and using data for day-to-day patient care provision in critical care environments means that the use of data for quality improvement seems not too big a step.

## Conclusion

Given data quality issues are systemic in many low- and middle-income countries, it is not surprising that difficulties were seen in maintaining quality data entry into the WHO TM in Rwanda over a 4-year implementation period. Despite enthusiasm for the registry, the desire to implement the 251 questions of WHO’s TM without modification may have let the perfect be the enemy of the good-enough. Indeed, after conducting this assessment, we limited the data collection in Rwanda to 52 variables; these are now collected with over 90% case and variable completeness – the threshold of ≥80% completeness and accuracy has now been determined to be good enough for local needs - although stipends for data collection and data-quality improvement cycles are used to incentivise performance. We believe lessons learnt in Rwanda’s implementation of the WHO TM have applicability for other registries implemented in other low-income countries and beyond.

## Acknowledgement

Funding for this study was provided by the National Institute of Health Research, United Kingdom, through NIHR grant no:133135 (NIHR Global Health Group on Equitable Access to Quality Health Care for Injured People in Four Low or Middle Income Countries: Equi-injury) and NIHR grant no: 203062 (Rwanda912: Use of an innovative electronic communications platform to improve pre-hospital transport of injured people in Rwanda). The funders were not involved in the development of the study design, implementation, data collection, analysis and interpretations, nor the writing of manuscript and decisions on submission of the paper for publication.

## Data availability statement

The dataset analysed during the current study are not publicly available due to patient data confidentiality and data sharing restrictions of the Ministry of Health of the Republic of Rwanda. However, analysed data can be made available upon reasonable request, with permission of the Ministry of Health of the Republic of Rwanda.

