## Supplementary material for "Data quality and associations over 4 years of use of WHO’s Clinical Registry - Trauma Module, in Rwanda"

### Supplementary materials

#### Appendix 1:

*Appendix Table 1 - Schedule of formal trainings conducted for capacity building of Trauma Module data collectors*

| No. | Times when training provided | Locations | Description of training | Resources and persons |
|---|---|---|---|---|
| 1 | June 2019 | Hospitals 1,2,3 & 4 | Training for initiation of data collection. The trainees were introduced to the dataset including the definitions of all variables and navigating the TM for data reporting. | On-site, in person trainings to selected data collectors by RBC staff. |
| 2 | August 2021 | Hospitals 2,4,5 & 6 | Training for the 2 new health facilities to initiate data collection. Additional training for 2 of the initial facilities who had recruited replacement data collectors. | On-site, In person training for data collectors by RBC staff. |
| 3 | September 2021 | All 6 hospitals | Online training on data collection including the data entry process, prior to initiation of WHO/AFEM funding. | Virtual training by WHO team member. |
| 4 | January 2022 | Hospitals 2,4,5 & 6 | Follow up training for the 2 new hospitals and 2 initial hospitals. Including field visits to review implementation of the TM; refresher training on data entry; and to identify gaps and challenges faced by data collectors. | On-site, In person training for data collectors by RBC staff. |
| 5 | May - June 2022 | All 6 hospitals | Training of injury registrars on injury data registration and the Abbreviated Injury Scale (AIS), towards building their capacity to abstract injury data from dashboards, rule out information that is not code-able and to distinguish between injuries and outcomes. Further training provided to familiarize the dashboards, enabling the interpretation and use of local | On-site, In person training for all data collectors by RBC Resource persons. |

| | | | | |
|---|---|---|---|---|
| | | | data. Challenges and gaps hindering the friendly use of the system as well as the interpretation of AIS that is generated by the system were reviewed. | |
| 6 | July 2022 | All 6 hospitals | Onsite training to all data collectors to improve the quality of data reported into TM. | On-site, In person training for all data collectors by RBC staff. |

### Appendix 2:

*Appendix Table 2 – Percentage Variables Missingness by Trauma Module domains*

| Injury Module domains | Median Variable missingness (%) | Total number of valid variables* | No. of variables with <25% data missingness (%) | No. of variables with >75% data missingness (%) |
|---|---|---|---|---|
| Registry & Demography | 27.57 | 20 | 10 (50.00%) | 7 (35.00%) |
| Pre-ED care | 63.34 | 20* | 9 (45.00%) | 10 (50.00%) |
| Context of the Injury | 29.65 | 31 | 15(48.39%) | 9 (29.03%) |
| Details of the Injury | 26.50 | 19 | 9 (47.37%) | 0 (0.00%) |
| Care in the ED | 44.15 | 27 | 13 (48.15%) | 7 (25.93%) |
| ED Disposition | 51.67 | 9* | 3 (3.33%) | 2 (22.22%) |
| In-ward Care & Facility Disposition | 77.30 | 22 | 7 (31.82%) | 12 (54.55%) |

* Valid variables are those available in the system for entry of data, based on the prior responses. If some earlier variables had nil responses, it would result in the non-generation of subsequent variables for data entry, due to skip patterns. As such, 3 variables (2 in Pre-ED care and 1 in ED Disposition domains) did not appear. ED refers to Emergency Department.
